# Social Determinants of Health and Functional Brain Connectivity Predict Long-Term Physical Activity in Older Adults with a New Cardiovascular Diagnosis

**DOI:** 10.1101/2024.09.30.24314678

**Authors:** Nagashree Thovinakere, Satrajit S. Ghosh, Yasser Itturia-Medina, Maiya R. Geddes

**Author notes:** Corresponding Author: Nagashree Thovinakere, The Neuro, Montreal Neurological Institute McGill University 3801 Rue University Montreal, QC, H3A 2B4.

## Abstract

**Background:** Physical activity is essential for preventing cognitive decline, stroke and dementia in older adults. A new cardiovascular diagnosis offers a critical window for positive lifestyle changes. However, sustaining physical activity behavior change remains challenging and the underlying mechanisms are poorly understood.

**Methods:** To identify the neural, behavioral and contextual predictors of successful longer-term behavior change after a new cardiovascular diagnosis, we used support vector machine learning to predict changes in moderate-to-vigorous physical activity over four years in 295 cognitively unimpaired older adults from the UK Biobank, testing three models that incorporated baseline: (i) demographic, cognitive, and contextual factors, (ii) baseline resting-state functional connectivity alone, and (iii) combined multimodal features across all predictors.

**Results:** The combined multi-modal model had the highest predictive power (r=0.28, p=0.001). Key predictors included greenspace access, social support, retirement status, executive function, and between-network functional connectivity within the default mode, frontoparietal control and salience/ventral attention networks.

**Conclusions:** These findings underscore the importance of social and structural determinants of health and uncover neural mechanisms that may support lifestyle modifications. In addition to furthering our understanding of the mechanisms underlying successful physical activity behavior change, these findings help to guide the design of interventions and health policy with the ultimate goal of preventing cardiovascular disease burden and late-life cognitive decline.

## 1. Introduction

Cardiovascular diseases substantially elevate the risk of dementia and stroke due to shared pathophysiological mechanisms across the heart–brain axis^1^. Indeed, mixed dementia, comprised of combined vascular and Alzheimer pathological changes, is the most prevalent etiology of dementia in older age^2^. With the global dementia burden projected to rise to 132 million by 2050^3^, there is an urgent need for targeted strategies to mitigate the vascular contributions to late-life cognitive decline. Physical activity is highly effective in lowering dementia risk and all-cause mortality among individuals with cardiovascular disease^4,5^. Thus, targeting physical activity engagement as a strategy for dementia prevention following a cardiovascular diagnosis, is essential^6,7,8,9^.

Despite the well-established benefits of physical activity, physical inactivity remains prevalent, with approximately 27.5% of the global population not meeting recommended activity levels^10^. The prevalence is high among older adults and has escalated since the COVID-19 pandemic, especially among older adults with chronic conditions^11–13^. Moreover, motivating and maintaining long-term behavior change is difficult. Observational studies report that only 4.3% of individuals adopt lifestyle modifications within six months following a cardiovascular event, with adherence rates dropping to 3–11% after five years^14^. Understanding why and when individuals engage in initiation of physical activity is crucial for designing effective interventions.

To move towards a precision medicine approach to behavior change, it is important to go beyond group-level statistical approaches to identify individual differences and contextual factors at the level of the individual^28,71^. Prior behavioral research applying group-level statistics has highlighted factors such as self-efficacy^17^, self-regulation^18^, and biological sex, where males generally show higher adherence rates than females^19^ in influencing physical activity engagement. Psychological factors, including depression, fatigue, and executive function have also previously been shown to influence adherence^20,21^. Furthermore, social and structural determinants of health, which refers to the environmental conditions in which individuals are born, live, learn, work, play, and age have a cumulative impact on physical, mental, and brain health^22,23^. Factors such as access to greenspace and neighborhood walkability^25^, social support^26^, socioeconomic status^24^ are strongly associated with physical activity levels. Critically however, whether these factors also support physical activity behavior change remains unknown. These determinants may not only shape physical activity behavior but also act as upstream contributors to disparities in health outcomes, including incidence of dementias^27^. Further, neuroimaging provides insights into individual differences in brain organization and highlights neurodiversity, that is, how brain functions vary across individuals based on multilevel factors non-modifiable factors (e.g., genetics, biological sex) and differential life exposures to social and structural determinants of health^29^.

Functional connectivity offers a promising avenue for characterizing complex brain-behavior relationships^72,73^. Predictive modeling based on functional connectivity^72^ leverages the most relevant features of functional connectivity to predict behavioral outcomes. By mapping the brain’s intricate connections and integrating them with data on individual behaviors, it offers a window into the neural basis of highly complex phenomena. Indeed, prior research supports the utility of functional brain connectivity for behavioral prediction^72,74^, and shows that it can outperform the predictive power of structural features for lifestyle adherence^76^. This is in line with the Stern theoretical framework of cognitive reserve (the ability to maintain function in the face of age- and disease-related brain changes) that suggests functional measures might best capture the “neural implementation” of cognitive reserve^77^.

To better understand the drivers of physical activity behavior change among older adults who stand most to benefit, the current study adopts a precision medicine framework combined with a whole-brain machine learning approach. Specifically, we examine the roles of sociodemographic factors (e.g., age, sex, socioeconomic status), behavioral characteristics (e.g., retirement status, general health, pain, depression), cognitive function (e.g., attention, executive function), social factors (e.g., networks and support), environmental context (e.g., access to green spaces), and baseline resting-state functional connectivity (RSFC) on future physical activity behavior change after physically inactive older individuals receive a new cardiovascular diagnosis. This comprehensive approach is designed to uncover tangible targets for future interventions, including public policy changes, tailored to individual needs for those at a heightened risk of cognitive decline. By employing a rigorous data-driven machine learning approach, the current study aims to uncover the neurobehavioral mechanisms driving successful physical activity at the individual level^29^.

## 2. Methods

### 2.1 Participants

295 (mean age = 63.13 years ± 7.5, 188 women) cognitively unimpaired and physically inactive older adults from the UK Biobank, a large-scale population-based longitudinal cohort were included in this study. Inclusion criteria were: 1) cognitively unimpaired at enrollment; 2) reported a new cardiovascular diagnosis (i.e., hypertension, type II diabetes, dyslipidemia, cardiac angina or myocardial infarction) between baseline (T1; 2014) and follow-up over four years later (T2; 2019) (mean duration 4.2 years, SD 1.1); 3) did not meet the World Health Organization recommendation of 150 minutes/week of moderate-to-vigorous physical activity (MVPA) at baseline^3^; and 4) age >= 60. These criteria yielded a final sample size of 295 after removing four participants for having poor quality brain imaging data. Unimpaired cognition was defined as follows: performance scores on each cognitive test were converted into a percentile rank, and the raw score corresponding to the 5^th^ percentile (or 95^th^, on tests where higher scores represented worse performance) was identified as the cut-off for impairment^54^. An illustration of the study timeline is shown in Fig. 1. The brain imaging visit (Instance 2 of the UK Biobank) was considered the baseline timepoint, and the first repeat imaging visit (Instance 3 of the UK Biobank) was considered the follow-up timepoint. Demographic variables including age, sex, years of education, household income, and socioeconomic status (as measured through Townsend deprivation index)^55^ were included as covariates of non-interest. Average total household income before tax was divided into five groups (< £18,000, £18,000 to 30,999, £31,000 to 51,999, £52,000 to 100,000, and > £100,000). MRI data were obtained at baseline, before participants had received a new cardiovascular diagnosis, and moderate-to-vigorous physical activity (MVPA) self-reported data and cognitive indices were obtained for the two time-points: at baseline and in follow up after 4 years (mean duration 4.2 years, SD 2.1; ranging from 8 months to 4.8 years). The distribution of cardiovascular conditions was as follows: 183 individuals with hypertension, 20 with diabetes, 161 with high cholesterol, and 10 with cardiac angina or myocardial infarction. Inventories used to measure physical activity, psychosocial, cognitive, and environmental factors in the current study are briefly described below. Please refer to the Supplementary materials for further details. Medication use was measured at follow-up. Medication use was prevalent in this cohort, with 141 individuals taking cholesterol-lowering medication, 162 taking blood pressure medication, and 85 using both. Participant demographic characteristics are summarized in Table 1.

**Fig. 1.**
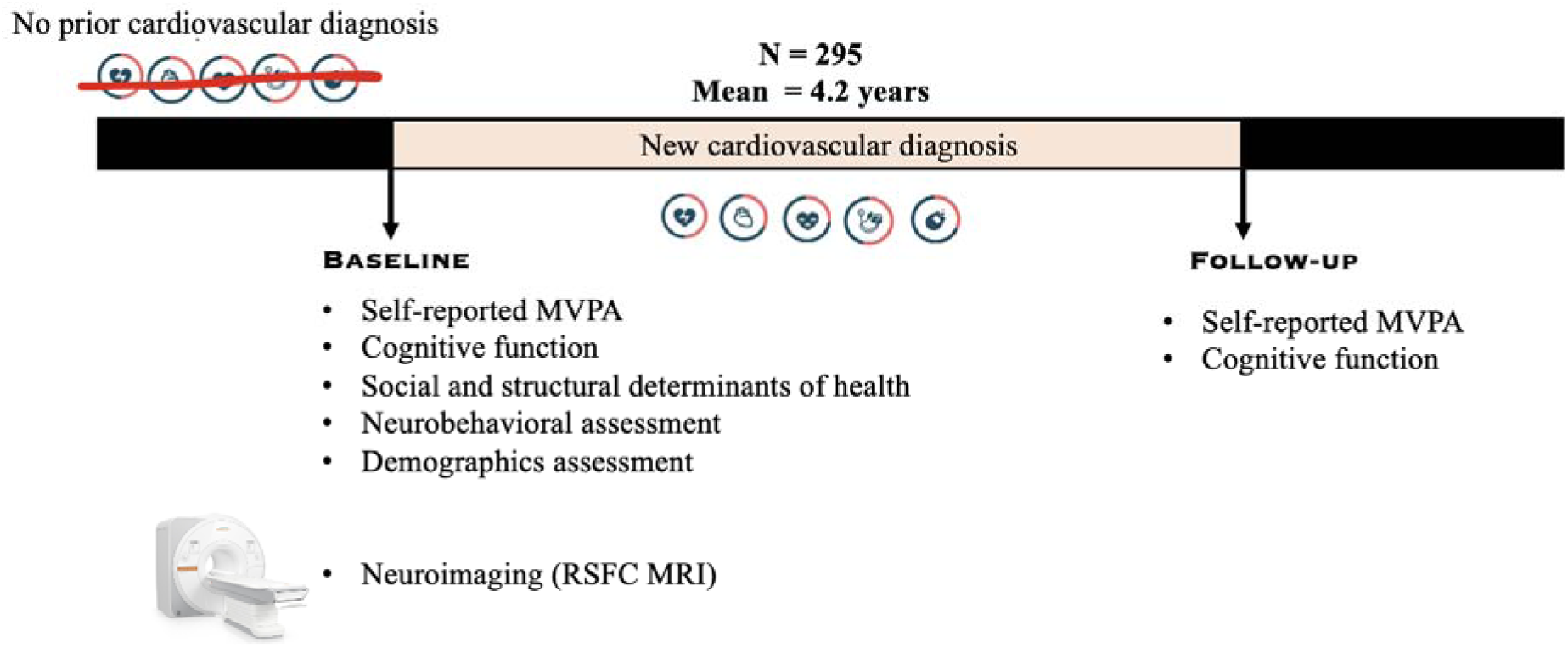
Study Timeline: Older participants received a new cardiovascular diagnosis (i.e., hypertension, type II diabetes, dyslipidemia, cardiac angina or myocardial infarction) between the baseline and follow-up periods (mean 4.2 years), with no cardiovascular diagnoses reported prior to baseline. Assessments included self-reported moderate-to-vigorous physical activity (MVPA), cognitive function, neurobehavioral factors (such as depression, anxiety, general or pain), social and structural determinants of health (including social support, retirement status and greenspace access). Resting-state functional connectivity (RSFC) MRI was assessed at baseline.

**Table 1:** Participant baseline demographic information for the UK Biobank sample. SD = Standard Deviation. MVPA = Moderate to Vigorous Physical Activity.

| Demographic Factor (n = 295) | Mean±SD |
| --- | --- |
| Age at baseline (years) | 63.13 years ± 7.5 |
| Female sex (count and %) | 188 women (63.72%) |
| Household income (£, most frequent range) | 18,000 to 30,999 (61%) |
| Education (years) | 15.4±3.2 |
| Townsend deprivation index score | -1.3±3.2 |
| MVPA at baseline (min/week) | 101.62±4.5 |
| MVPA at follow-up (min/week) | 109.10±6.8 |
| Frequency of friends and family visits at baseline | 4.2±1.03 |
| Able to confide at baseline | 3.8±1.67 |
| Greenspace proximity at baseline (%) | 34.1±20.59 |
| Coastal proximity at baseline (%) | 0.89±2.88 |
| Depression score at baseline | 2.1±1.02 |
| Anxiety score at baseline | 1.8±0.88 |
| Number retired at baseline (count and %) | 111 retired (37.64%) |

### 2.2 Data analysis overview of behavioral and contextual factors

Participants completed a comprehensive battery of psychosocial, behavioral, cognitive, and environmental assessments at both baseline and follow-up (Fig. 1). These assessments are briefly outlined below. To investigate the relationship between physical activity and cognition, we first examined whether baseline cognitive function predicted future physical activity behavior. We then assessed whether increases in physical activity at follow-up were linked to cognitive gains. Next, we evaluated whether social and structural health determinants at baseline predicted successful future engagement in physical activity. Resting state functional connectivity (RSFC) was assessed at baseline only, prior to any cardiovascular diagnosis, thereby reducing potential confounding effects related to blood flow alterations^56,57^ (Makedonov et al., 2013; Tsvetanov et al., 2021). A full list of input variables used in the prediction models is available in Supplementary Table 1.

#### 2.2.1 Townsend deprivation index

The Townsend Deprivation Score is an area-based score of social deprivation aggregated from percentage of unemployment rate, non-car ownership rate, non-home ownership rate and household overcrowding (proportion of households with more people than rooms). This indicator was determined immediately prior to the participant joining the Biobank and was based on data from the preceding national census^58^. The Townsend Deprivation Index is a composite, standardized score with higher positive values indicating greater socioeconomic deprivation and lower (negative) values indicating less deprivation. Each participant was assigned a score corresponding to their postal code area.

#### 2.2.2 Physical activity questionnaires

Successful future physical activity engagement, the primary behavioral outcome of interest, was defined as the difference between the overall MVPA in minutes per week measured at follow-up compared to the overall MVPA in minutes per week measured at baseline. This change in MVPA was assessed using the Lifetime Total Physical Activity Questionnaire^59^, which captures self-reported MVPA by recording the frequency and duration of each physical activity type performed weekly. The total time spent on moderate and vigorous activities was then calculated to derive the overall MVPA in minutes per week. This total score served as an indicator of each individual’s physical activity engagement. The scale was administered at both baseline and follow-up timepoints to assess changes over time.

Leisure-time physical activity was also measured through items capturing activities such as walking for pleasure, light and heavy do-it-yourself (DIY) tasks (e.g., pruning, watering the lawn, carpentry, digging, weeding), and recreational activities (e.g., swimming, cycling, bowling). The total time spent on these activities was then calculated to derive the overall leisure time physical activity in minutes per week.

Occupational physical activity was assessed with questions adapted from the UK Biobank, including “Does your work involve heavy manual or physical work?” and “Does your work involve walking or standing for most of the time?” These questions helped capture physical activity levels related to participants’ work environments. Scores ranged from 1 (Never/rarely) to 4 (Always) and were treated as a continuous measure.

#### 2.2.3 Cognitive assessments

A computerized cognitive battery was administered using a touchscreen tablet. The tests were specifically developed for the UK Biobank and have been validated^54^, while sharing features with established cognitive assessments. The battery included the following tasks: Reaction time, Numeric memory, Prospective Memory, Fluid intelligence, Matrix pattern completion, Tower rearranging, and Trail making. A detailed description of these tasks can be found in Supplementary materials Appendix A.

#### 2.2.4 Social support

The measures available in the UK Biobank for social support come from the items “How often do you visit friends or family or have them visit you?” and “How often are you able to confide in someone close to you?” Participants rated each item on a Likert scale from 0 (Never or almost never) to 6 (Almost daily). For the frequency of visits, the categories “never or almost never” and “no friends or family outside the household” were combined into a single category, “never.” This adjustment was made because these responses were similar, and there were only a few participants with no friends or family outside the household (n = 16). Scores ranged from 0 to 6 and were treated as a continuous measure. Loneliness was also assessed using the item “Do you often feel lonely?”. Responses were recorded as yes (1) or no (0).

#### 2.2.5 Greenspace and coastal proximity assessment

Environmental indicators included in this study were the proportion of green space and water within 300 m of residential addresses, using the 2005 Generalised Land Use Database for England and Centre for Ecology and Hydrology 2007 Land Cover Map data for Great Britain^60^. The buffer size of 300m was decided based on relevant health evidence and public policy on both density and accessibility. Coastal proximity was estimated using Euclidean distance raster^61^.

#### 2.2.6 Psychosocial and mental health factors

Psychosocial factors were assessed through self-reported experiences, including depression, anxiety, general pain, and lifestyle factors such as retirement status. Depression was evaluated using two items: “Feeling down, depressed, or hopeless” and “Little interest or pleasure in doing things.” Participants rated their experiences on a four-point scale, ranging from 0 (Not at all) to 4 (Nearly every day). Anxiety was assessed similarly, with two items: “Feeling nervous, anxious, or on edge” and “Not being able to stop or control worrying.” General pain was measured using a single item: “Have you had pains all over your body for more than 3 months?” Responses were recorded as yes (1) or no (0). Participants also rated their overall health perception on a scale from 1 (Excellent) to 4 (Poor). Additionally, participants indicated their retirement status with a simple yes (1) or no (0) response. These assessments were conducted at both baseline and follow-up timepoints.

### 2.3 MRI Data Acquisition

Details of image acquisition and processing are available in the UK Biobank Protocol (http://biobank.ctsu.ox.ac.uk/crystal/refer.cgi?id=2367), and Brain Imaging Documentation (http://biobank.ctsu.ox.ac.uk/crystal/refer.cgi?id=1977). Briefly, all brain MRI data were acquired on a Siemens Skyra 3T scanner with a standard Siemens 32-channel RF receiver head coil, using the following parameters: TR = 2000 ms; TI = 800 ms; R = 2; FOV = 208 × 256 × 256 mm; voxel size = 1 × 1 × 1 mm. For resting-state fMRI scans, two consecutive functional T2*-weighted runs were collected with eyes closed using a blood oxygen level dependent (BOLD) sensitive, single-shot echo planar imaging (EPI) sequence with the following parameters: TR = 735 ms; TE = 39 ms; flip angle = 52°; FOV 88 x 88 x 64 matrix; resolution = × 2.4 × 2.4 mm; 490 volumes; and acquisition time = 6 minutes per run.

### 2.4 Resting-State Functional MRI Data Preprocessing

Preprocessing of raw functional images from the UK Biobank was done using the fMRIprep pipeline (version 20.2.4)^62^. For each of the BOLD runs per participant, the following preprocessing was performed: First, the T1w reference was skull-stripped using a Nipype implementation of the antsBrainExtraction.sh (ANTs) tool. A B0-nonuniformity map (or fieldmap) was estimated based on a phase-difference map calculated with a dual-echo gradient-recall echo (GRE) sequence, which was then co-registered to the target EPI reference run and converted to a displacements field map. A distortion-corrected BOLD EPI reference image was constructed and registered to the T1-weighted reference using a boundary-based approach (using bbregister, Freesurfer). Rigid-body head-motion parameters with respect to the BOLD EPI reference were estimated (using mcflirt, FSL 5.0.9)^63^ before spatiotemporal filtering was performed. BOLD runs belonging to the single band acquisition sessions were slice-time corrected (using 3dTshift, AFNI 20160207). The BOLD time series were resampled into their original, native space by applying a single, composite transform to correct for scan-to-scan head motion and susceptibility distortions. Functional scans were spatially smoothed using a 6 mm full width at half maximum Gaussian smoothing kernel.

Additional preprocessing steps were undertaken to remove physiological, subject motion, and outlier-related artifacts, which were implemented using the nilearn package. Non-neuronal sources of noise from white matter and CSF were estimated and removed using the anatomical CompCor method^64^ to allow for valid identification of correlated and anticorrelated networks^65,66^. Temporal band-pass filtering (0.008–0.09 Hz) was then applied. Additionally, scan-to-scan mean head motion (framewise displacement) was used as a covariate of non-interest in all second-level analyses (mean head motion = 0.2 mm, SD = 0.1 mm). Head motion is a known important potential confound as it produces systematic and spurious patterns in connectivity and is accentuated in Alzheimer’s disease (AD) and cognitively typical aging populations^67^. Critically, we did not identify a relationship between the mean head motion parameter and the primary behavioral variable of interest, physical activity change (all p > 0.05). The framewise displacement timeseries was determined by calculating the maximum shift in the position of six control points situated at the center of a bounding box around the brain, computed independently for each scan. Four participants were removed from the UK Biobank sample final analysis for having >30 scan volumes flagged, leading to the final sample size of 295 participants. This cut off was determined based on preserving at least 5 minutes of scanning time^68^.

### 2.5 Machine Learning Modelling

To predict future successful physical activity (MVPA) behavior change as a continuous measure following a new cardiovascular diagnosis, we used the support vector machine (SVM) algorithm from the scikit-learn (v0.21.3) library, utilizing the pydra-ml (v0.3.1) toolbox. Three separate models were trained: (1) combined demographic, cognitive and contextual features only (2) neuroimaging features only, and (3) multimodal model combining all demographic, cognitive, contextual and neuroimaging features. Contextual features encompass many factors influencing responses to interventions and overall clinical outcomes, including but not limited to the personal characteristics, and social and structural determinants of health^14,19,23^. This multilevel, complexly interacting framework is essential for understanding physical activity behavior change in individuals with cardiovascular disease and optimizing the effectiveness of preventive strategies and interventions.

The SVR works by placing constraints to ensure only a small number of observations (support vectors) are used. SVR works with the goal of constructing a regression line that fits the data within some chosen level of error. We used the default parameters, which include the radial basis function kernel to capture non-linearities in the data. To assess the robustness of our findings, we repeated our analysis using additional machine learning algorithms of increasing complexity, defined by the computational resources required for model simulation. Specifically, we examined linear regression, random forest, and multi-layer perceptron algorithms, using default parameters unless otherwise specified. Further details on these algorithms are available in the supplementary materials Appendix B.

We investigated model performance using four features selection strategies for all the prediction models we tested (Linear Regression, SVM regression, Random Forest regression, and multi-layer perceptron): (1) using all features, (2) removing redundant neuroimaging features, (3) selecting only the top 20 features, and (4) excluding the top 20 features to assess their necessity for predictive performance. To generate independent test and train data splits, we used a bootstrapped group shuffle split sampling scheme. For each iteration of bootstrapping, a random selection of 20% of the participants, balanced between the two groups, was designated as the held-out test set. The remaining 80% of participants were used for training. This process was repeated 50 times, fitting and testing the four classifiers for each test/train split. We used the default of 50 bootstrapping splits from pydra-ml toolbox. We provide several interpretable measures of model performance based on the observed vs predicted values; Pearson’s r correlation, the squared correlation, R^2^, root mean squared error (RMSE), which measures the average prediction error as the average difference between the observed and predicted values and the mean absolute error (MAE) as the average absolute difference between the observed and predicted values. RMSE and MAE are related with MAE being less sensitive to outliers and the lower the value the better the model performance. The p-value for each model is derived by comparing the correlation coefficient between the observed and predicted values to a null distribution derived from 1000 non-parametric permutations. Age, sex, years of education, and medication use were controlled as covariates in all prediction models.

We employed Kernel SHAP (SHapley Additive exPlanations)^69^ to assess the significance of baseline RSFC features in predicting successful engagement in physical activity. We computed the average absolute SHAP values across all predictions, weighted by the model’s median performance, and calculated mean SHAP values across splits for each model. This entire pipeline, encompassing machine learning models, bootstrapping, and SHAP analysis, was implemented using pydra-ml toolbox.

### 2.6 Reducing collinearity using Independence Factor to enhance model interpretability

Collinearity among features can significantly affect model generation and interpretation, particularly in resting state fMRI - analyses. To address this, we employed the Independence Factor method^69^, which iteratively removes features with strong dependence above a set threshold, ensuring a consistent set of features across models. Using distance correlation, which accommodates non-monotonic relationships, we systematically increased the threshold to eliminate redundant features while preserving model performance within a narrow margin. Importantly, reducing distance correlation enhances statistical independence among features, thereby improving model interpretability. We applied thresholds ranging from 1.0 (keeping all features) to 0.2 (removing features with distance correlation above 0.2). Our goal was to identify a feature set that maintained model performance within three percentage points of using all features, resulting in a more parsimonious and interpretable model without compromising accuracy, essential for clinical applicability.

### 2.7 Performance using most important and least important features

To address the question of why certain features are important, we evaluated model performance under two scenarios: one using only the top 20 features and another excluding these features. This method mitigates the common pitfall in brain-behavior prediction analyses, where the significance of the top features may not reflect their true impact on model performance. By comparing performance metrics in both scenarios, we can gain a more nuanced understanding of the highlighted features’ contributions and derive mechanistic insights into the neural correlates of successful behavior change.

This article was prepared according to the guidelines outlined in TRIPOD + AI statement: updated guidance for reporting clinical prediction models that use regression or machine learning methods^83^. The checklist is available in supplementary materials.

## 3. Results

### 3.1. Behavioral Results

Following a new cardiovascular diagnosis, participants demonstrated a significant average increase in physical activity engagement of 7.48 min/week ± 1.23, reflecting a 7.36% increase in moderate-to-vigorous physical activity (MVPA) (r=0.38. p < 0.01). A positive trend was observed between higher baseline MVPA and change in MVPA at follow-up among inactive older adults (r = 0.51, p = 0.12). No significant associations were identified between medication use (cholesterol-lowering or blood pressure) and either baseline MVPA or change in MVPA following a new cardiovascular diagnosis. Baseline cognitive function across multiple domains, including processing speed and executive function, was not significantly associated with baseline MVPA. Moreover, changes in cognitive function (i.e., follow-up minus baseline scores/baseline) were not associated with either change in MVPA or baseline MVPA.

### 3.2. Prediction Modeling Results

Prediction of future change in physical activity (MVPA as a continuous variable) among 295 cognitively unimpaired older adults, was conducted separately across three support vector machine (SVM) learning models with inputs that included baseline neuroimaging, behavioral or combined features as predictors: (Model 1) demographic, cognitive, and contextual features, (Model 2) RSFC MRI inputs, and (Model 3) a multimodal model integrating all behavioral and neural features. As shown in Table 2, the model based solely on demographic, cognitive, and contextual features did not significantly predict changes in MVPA at follow-up (r=0.17, p=0.056). In contrast, the neuroimaging model (r=0.25, p=0.004, FDR-corrected) and the multimodal model combining all features (r=0.28, p=0.001, FDR-corrected) significantly predicted MVPA change. SVM models consistently outperformed other machine learning algorithms, including linear regression, random forest, and multi-layer perceptron (performance metrics for the other algorithms described in Supplementary Table 2).

**Table 2:** Performance metrics derived from SVM regression models. R and R^2^ represent the Pearson correlation and the squared correlation between the predicted and observed values, respectively. Root mean squared error (RMSE) represents the average difference between the observed and predicted values (average prediction error). Mean squared error (MAE) represents the absolute mean difference between the predicted and observed values. SSDoH represents the social and structural determinants of health.

| Model | r | R<sup>2</sup> | RMSE | MAE | p-value |
| --- | --- | --- | --- | --- | --- |
| Behavioral and SSDoH | 0.17 | 0.02 | 0.13 | 0.11 | 0.058 |
| Neuroimaging | 0.25 | 0.07 | 0.11 | 0.09 | 0.004 |
| Multimodal | 0.28 | 0.08 | 0.11 | 0.09 | 0.001 |

A predictive model that generalizes to different settings has greater clinical utility than a model that only works under specific conditions. The SVM model demonstrated robust performance across all scenarios (Supplementary Table 3). Given the high dimensionality of resting state fMRI data, Independence Factor Analysis^69^ was applied to neuroimaging features, resulting in an optimal subset of 250 features for subsequent analyses. After removing highly dependent features, based on distance correlation, from the original 400 neuroimaging features, the final model included 250 neuroimaging features and 19 demographic, cognitive, and contextual features, for a total of 269 features.

Mean SHAP (SHapley Additive exPlanations) values illustrating feature importance across the three models are summarized in supplementary table 4. In the multimodal model, neighborhood greenspace percentage, social support (i.e., frequency of visits from friends and family), retirement status, and occupational physical activity showed a significantly positive association with MVPA change, indicating that higher greenspace exposure, more frequent friend and family visits, not being retired, and greater occupational physical activity predicted greater improvements in MVPA (p < 0.05, FDR-corrected). For cognitive features, improved higher executive function (the Tower Rearranging task) emerged as a significant predictor of future increase in MVPA (supplementary Table 4), while no other behavioral, cognitive, or contextual features showed significant prediction effects.

Figure 2 highlights the most significant baseline RSFC MRI features from the highest-performing multimodal prediction model. These features were primarily located within the left hemisphere and spanned multiple large-scale networks, with critical nodes in the default mode network (e.g., temporal lobe and medial prefrontal cortex), frontoparietal control network (e.g., lateral prefrontal cortex), and salience/ventral attention network (e.g., frontal operculum) (Figure 2b). Of the top RSFC nodes, 7 were within the default network, 7 were within the frontoparietal control network, 6 were within the salience ventral attention network. Enhanced RSFC within the default mode network was associated with increased physical activity at follow-up. Moreover, increased MVPA was associated with greater positive RSFC between frontoparietal control network and the default mode network.

**Figure 2.**
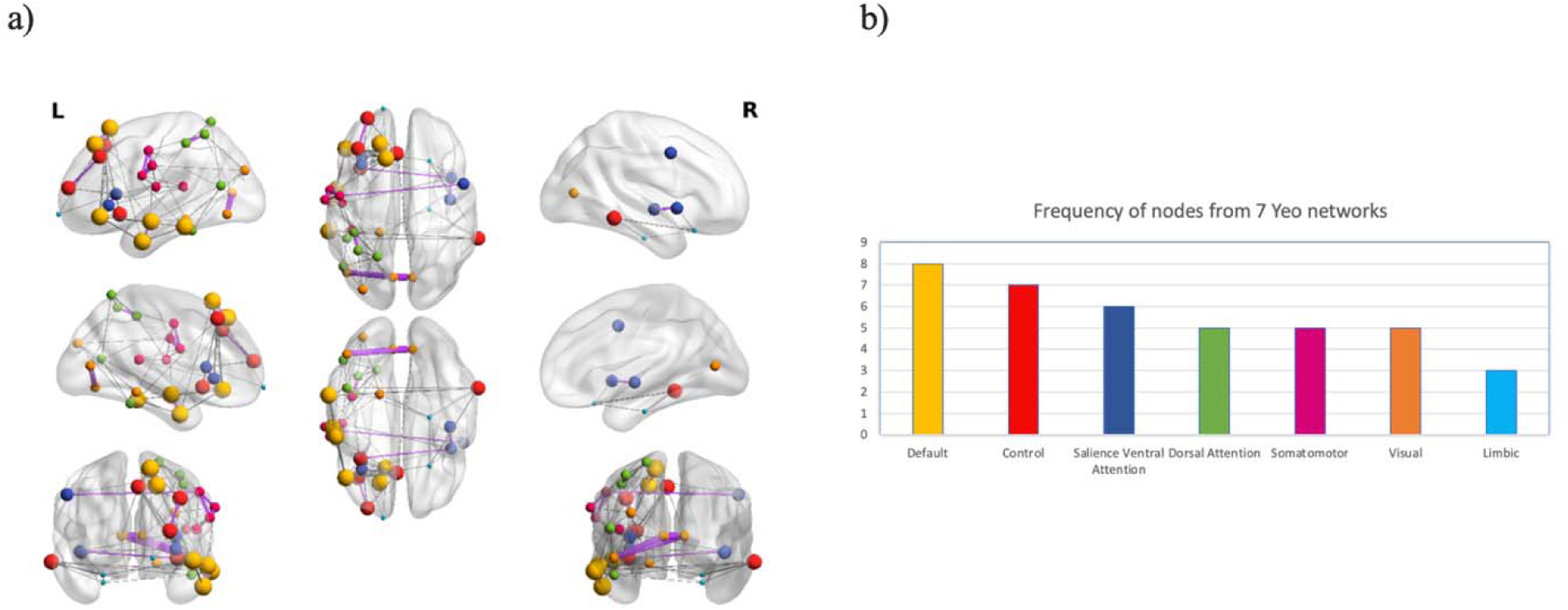
Baseline RSFC features that predict future increase in MVPA after a new cardiovascular diagnosis in aging. (a) Neuroanatomical depiction of significant features from the multimodal model and their corresponding importance values: Node size (spheres) depicts the frequency of that brain region among predictive features, while edge thickness (line connecting two nodes) represents the weight or importance of a predictive RSFC feature. Purple signifies positive RSFC whereas grey signifies negative RSFC associated with enhanced physical activity at follow up compared to baseline. (b) The summary of frequency and distribution of predictive nodes grouped by location within canonical neural networks (i.e., Yeo 7 networks).

## 4. Discussion

In this study, we systematically evaluated whether neural, cognitive, behavioral or social and structural determinants of health (SSDoH) predict successful long-term physical activity behavior change among inactive older adults newly diagnosed with a cardiovascular illness. Our findings highlight the importance of SSDoH, particularly access to greenspace, social support, occupational physical activity and retirement as key predictors of positive changes in physical activity (i.e., MVPA) following cardiovascular disease diagnosis in aging. From a cognitive standpoint, improved performance on the tower rearranging task, a measure of executive function (i.e., goal-directed planning) was significantly associated with positive physical activity behavior change. We found that a multimodal model incorporating behavioral, contextual, and neuroimaging features provided the strongest predictive value. Functional connectivity analyse revealed that sustained increases in MVPA were linked to greater within-network connectivity in key regions of the default mode network and enhanced between-network connectivity between the default mode and frontoparietal networks. These predominantly left-lateralized connections localized primarily within heteromodal cortices, underscoring the role of large-scale brain networks in facilitating behavior change.

The critical windows theory suggests that successful behavior change may be facilitated by an external threat from a major life event or circumstance (e.g., receiving a diagnosis of a new chronic illness such as a cardiovascular disease, pregnancy, or menopause), which might catalyze the reassessment of goals and increase motivation for change presenting a ‘teachable moment’ in life^15^. For example, individuals with chronic conditions, including diabetes and other cardiovascular diseases are often more likely to maintain or increase their leisure-time physical activity levels^16^. Consistent with this, our study observed increased physical activity behavior among older adults who reported a new cardiovascular diagnosis. Thus, life transitions may serve as critical windows for intervention, offering opportunities to promote long-term physical activity engagement.

Our findings build on the growing body of literature demonstrating the influence of SSDoH on age-related health outcomes; for example, the influence of upstream factors on downstream protective behaviors such as physical activity engagement. Consistent with prior research, proximity to greenspace and social support were linked to increased physical activity behavior change^25, 33^. Similarly, high social support from friends and family was significantly associated with enhanced MVPA^23,28^. However, we found that quantitative aspects of social support (e.g., frequency of visits from family and friends) were stronger predictors of behavior change than qualitative aspects (e.g., ability to confide in others or perceived loneliness). There is likely a complex, bidirectional relationship between social contact frequency and emotional support in influencing physical activity^34^.

Contrary to prior research suggesting that retirement can increase leisure-time physical activity^35^, we observed a decline in physical activity over a five-year follow-up period after retirement. This reduction may be partially attributed to diminished social interactions post-retirement. Furthermore, catalysts for retirement, such as health issues or caregiving responsibilities can impact an individual’s motivation, financial capacity, and physical ability to remain active^36^. Indeed, retirement due to disability is associated with a decline in physical activity levels^35^. This finding highlights the importance of life milestones (e.g., parenthood, death of a loved one) as critical windows for behavior change and potential opportunities for dementia prevention^70^.

Even modest increases in MVPA can yield substantial health benefits for individuals with cardiovascular risk factors^32^. However, comorbid conditions may necessitate personalized activity targets due to variability in clinically meaningful responses. By identifying individual differences in key factors influencing long-term behavior change, spanning behavioral, cognitive, neural, social, and structural determinants, our findings contribute to the growing evidence base that can be leveraged to develop scalable and effective personalized physical activity interventions. Despite mixed prior findings suggesting that antihypertensives and cholesterol lowering medications such as beta-blockers and statins can impair exercise capacity due to muscle fatigue or reduced endurance^32^, we did not identify a relationship between medication use and behavior change, suggesting these medications may not limit long-term MVPA engagement.

We identified neural markers that predicted successful physical activity behavior change among older adults following a cardiovascular risk diagnosis. Future increases in physical activity were associated with enhanced positive functional connectivity between the default mode network and frontoparietal network, as well as greater within-network connectivity in the default mode network. These findings align with prior research showing that network connectivity of regions within the default mode network, especially the prefrontal cortex, support compensatory mechanisms in aging^73,74^. Prior age-related neuroimaging research has shown that default mode network is associated with complex decision-making processes critical for adaptive behavior in aging^37–39^. Moreover, our finding of increased default mode to frontoparietal network coupling with enhanced physical activity behavior change supports the default-executive coupling hypothesis of aging^37–47^: This model suggests that goal-directed cognition in older adults increasingly relies on accumulated knowledge (semanticization of cognition) to offset declining cognitive control resources for successful behavior^37,80^. Default-executive coupling has been associated with positive behavioral outcomes including creative problem solving^78^ and autobiographical memory^37,79^. Our findings point to a possible large-scale network connectivity fingerprint as a marker of resilient aging and of individuals who may be the most receptive to changing their lifestyle behavior.

Finally, our observation that combining multimodal brain and behavioral features leads to an increase in model performance suggests that these features provide independent and relevant information for predicting changes in physical activity. Previous studies^48,49^ have also demonstrated that multimodal prediction models outperform unimodal ones. This improvement in prediction performance may arise because individual features capture distinct aspects of complex behaviors related to physical activity behavior change—insights that unimodal features alone may fail to capture.

Despite these contributions, several limitations of this work should be noted. First, self-reported measures of MVPA were used rather than objectively-measured physical activity measured using wearables. This choice was made due to the availability of accelerometry data at only one of the timepoints, making it impossible to measure behavior change. Self-reports should be interpreted with caution due to potential reverse causation effects, and significant variance between objectively measures and self-reported estimates of physical activity^82^. Second, objective measures such as accelerometers can differentiate between sedentary behavior, light activity, and moderate/vigorous activity, and can also provide physiological metrics for estimating cardiorespiratory fitness^50^. Finally, the correlational nature of functional connectivity analyses prevents us from determining causality in the brain behavior relationship identified^51,52,53^.

Nonetheless, our study has several notable strengths. It represents the largest and most comprehensive assessment of the brain, behavioral and contextual factors predicting successful longer-term physical behavior change after cardiovascular diagnosis in aging. This study highlights the importance of going beyond individual-level factors and considering structural factors such as greenspace and social support to promote physical activity behavior change, evidence that is critical to guide policy decision-making and urban planning. Future research must adopt a life course perspective to identify factors in younger or midlife adults and build a comprehensive understanding of physical activity behavior change across the lifespan.

## 5. Conclusion

This study demonstrated that individual differences in brain, cognition, behavior, and contextual factors, including social and structural determinants of health, drive a complex human behavior: Future engagement in physical activity among older adults that are newly diagnosed with a cardiovascular illness. Leveraging mechanistic predictors of future physical activity and adopting a precision medicine framework will potentially lead to targeted interventions that result in sustained behavioral change and dementia prevention.

## 6. Data and Code Availability

The individual-level UK Biobank data can be obtained from https://www.ukbiobank.ac.uk/. The code required to run the analyses is available through Github (https://github.com/nagatv11/cvd_MVPA.git).

## 7. Ethics Statement

This study utilized data from the UK-Biobank study, which obtained ethics approval from the Northwest Multi-Centre Research Ethics Committee (MREC, approval number: 11/NW/0382), and obtained written informed consent from all participants prior to the study. This research has been conducted using the UK Biobank Resource under Application No. 45551.

## Acknowledgements

We would like to thank Steven Grover and Michael Petrides for their helpful comments on this manuscript. This research used the NeuroHub infrastructure and was undertaken thanks in part to funding from the Canada First Research Excellence Fund, awarded through the Healthy Brains, Healthy Lives initiative at McGill University. This research was enabled in part by support provided by Calcul Québec and the Digital Research Alliance of Canada. This research was undertaken thanks in part to funding from a National Sciences and Engineering Research Council of Canada (NSERC) Discovery Grant (DGECR-2022-00299), an NSERC Early Career Researcher Supplement (RGPIN-2022-04496), a Fonds de Recherche Santé Québec (FRSQ) Salary Award, the Canada Brain Research Fund (CBRF), an innovative arrangement between the Government of Canada (through Health Canada) and Brain Canada Foundation, a Brain Canada Future Leaders Award, an Alzheimer Society Research Program (ASRP) New Investigator Grant, the Canadian Institutes of Health Research, the Canada First Research Excellence Fund, awarded through the Healthy Brains, Healthy Lives initiative at McGill University, and the National Institutes of Health (P30 AG048785) to MRG, and the Consortium pour l’Identification précoce de la Maladie d’Alzheimer-Québec (CIMA-Q) awarded to NT. SSG was partially supported by NIH projects P41EB019936 and RF1MH121885.

